# Unsupervised Machine Learning of Computed Tomography Angiography Features Uncovers Unique Subphenotypes of Aortic Stenosis With Differential Risks of Conduction Disturbances Following Transcatheter Aortic Valve Replacement

**DOI:** 10.64898/2026.02.24.26346951

**Authors:** Mustapha El Zeini, Madison Fang, Matthew P. Tran, Umarani Badarabandi, Chun Liu, Sachin B. Malik, Guson Kang, Nazish Sayed, Karim Sallam, Andrew Y. Chang, Ian Y. Chen

## Abstract

**Background:** Various measurements around the aortic valve are typically made on computed tomography angiograms (CTAs) before transcathether aortic valve replacement (TAVR) for aortic stenosis (AS), but their collective prognostic inference on periprocedural conduction disturbances (CDs) is not known. Here, we aimed to use unsupervised machine learning (UML) to analyze a multitude of pre-TAVR CTA features and uncover patient subphenotypes with differential risks of CDs.

**Methods:** Twelve nonredundant features involving the aortic valve, aortic root, and ascending aorta were extracted from the CTAs of 660 AS patients. UML of these features using agglomerative hierarchical clustering was performed on separate male and female datasets, with the optimal number of clusters determined by 30 cluster indices. Multivariable logistic regression was conducted to assess the dependence of CDs on cluster type and the latter’s incremental prognostic value over conventional risk factors.

**Results:** Three male clusters were optimally identified (M1-M3): M1 was associated with small valve leaflet calcification loads and aortic root dimensions; both M2 and M3 were associated with large valve leaflet calcification loads and a wide aortic root, but the aortic root was shorter in M2 than M3. Two female clusters were optimally determined (F1-F2): F2 was associated with larger valve leaflet calcification loads and aortic root dimensions. By logistic regression analysis, compared to M1 (reference), M2, but not M3, was more associated with CDs (OR_M2/M1_=2.15, *P*=0.032; OR_M3/M1_=2.12, *P*=0.085), with no difference between M3 and M2 (OR_M3/M2_=0.986, *P*=0.974) or between F1 and F2 (OR_F2/F1_=1.294, *P*=0.581). Including cluster type as a predictor in a regression model of CDs containing conventional risk factors as covariates improved the goodness-of-fit (*P*=0.020).

**Conclusions:** UML of pre-TAVR CTAs can reveal subgroups of male patients with differential risks for CDs and improve prognostication over conventional risk factors. UML-augmented pre-TAVR CTAs may help better guide personalized strategies to minimize CDs.

## INTRODUCTION

The initial introduction and later refinement of the transcatheter aortic valve replacement (TAVR) technique over the past decade have provided an effective life-prolonging treatment for patients with clinically significant aortic stenosis (AS).^1^ The widespread use of computed tomography angiography (CTA) for preoperative planning and advances in the transcatheter heart valve (THV) design have further helped to reduce many periprocedural complications, including perivalvular leak, conduction disturbances (CDs), coronary obstruction, and annular rupture.^2^ Among these, CDs persistently remain a major challenge to safe TAVR.^3^

Various predictors of CDs—including new onset left bundle branch block (LBBB) and high-degree atrioventricular block (HAVB) requiring permanent pacemaker implication (PPI)—have been examined, with THV type,^4^ implantation depth,^5^ and baseline right bundle branch block (RBBB)^6^ being the most well-validated. Additionally, specific locations of valvular/perivalvular calcification^6^ and membranous septal length^7^ measured on CTA have also been individually linked to altered risks of CDs. However, because TAVR involves fine dexterous manipulation of the delivery system and THV through the ascending aorta and the aortic root, we hypothesized that patient-specific anatomy centering around the aortic root—typically assessed as a multitude of dimensional measurements on preoperative CTA—may also impact CD risks.

The purpose of this study was to determine the extent to which various pre-TAVR CTA measurements around the aortic root collectively contribute to TAVR-related CDs. Here, we performed unsupervised machine learning (UML) in the form of agglomerative hierarchical clustering (AHC) to analyze a myriad of pre-TAVR CTA features and uncover specific subphenotypes of AS with differential risks for CDs. The knowledge obtained from this study should assist with the future incorporation of machine learning algorithms in pre-TAVR CTAs to enhance its prognostic capabilities.

## METHODS

### Data availability

The data that support the findings of this study are available from the corresponding author upon reasonable request.

### Enrollment of study participants

This retrospective study was conducted at both Stanford University and Veterans Affairs Palo Alto Health Care System (VAPAHCS) with prior approval and waiver of informed consent from the Institutional Review Board. The study was designed to eventually analyze the medical records of 660 patients, equally split among men and women. To achieve this, we retrospectively reviewed and screened the medical records of 1,046 patients (n=543 male and n=503 female) with clinically significant AS undergoing TAVR evaluations at Stanford University (October 2014 to March 2020) and VAPAHCS (July 2012 to January 2021). After applying predefined inclusion and exclusion criteria (**Table S1**), focusing on patients with degenerative calcific AS, native tri-leaflet aortic valves, and adequate pre-TAVR CTA images, we derived at a final cohort of 660 patients (n=330 each for men and women) for further analyses (**Figure S1**).

### Pre-TAVR CTA

Pre-TAVR CTAs were performed on either a GE Lightspeed VCT scanner (GE Healthcare; VAPAHCS only) or a Siemens SOMATOM Force CT System (Siemans Healthineers AG; both institutions) per clinical routines. Retrospective electrocardiography-gated image acquisition covering the aortic valve and aortic structures was performed after intravenous administration of iodinated contrast (Omnipaque 350, GE Healthcare; 4-5 mL/second for 30 seconds), followed by saline (80-100 mL at the same rate as contrast). A standard-pitch non-gated (GE) or high-pitched electrocardiography-triggered (Siemens) acquisition of the chest, abdomen, and pelvis was then followed to assess for vascular access. X-ray tube energy was either fixed at 120 kilovoltage peak (kVp) (GE) or automatically selected based on the scanner’s built-in algorithm (70-120 kVp; Siemens, CARE kV). Tube current modulation was performed from 30-70% of the RR interval at the discretion of the radiologist. Images were reconstructed at a slice thickness of either 0.625 mm (GE) or 0.750 mm (Siemens) for assessment of aortic root/ascending aorta dimensions, or 2.5 mm for quantification of aortic valve leaflet calcification loads.

### CTA image analysis and feature extraction

Reconstructed CTA images were analyzed using Aquarius iNtuition Ver.4.4.13.P3A (TeraRecon). Twenty-one common pre-TAVR CTA imaging features were initially extracted from patients’ CTA images according to clinical guidelines^8^ except for the calcification loads of individual valve leaflets, which were assessed as previously described (**Figure S2A**).^9^ This set of imaging features was further reduced to 12 features by trimming those highly correlated features (i.e., Pearson correlation coefficient ≥ 0.8). The resulting non-redundant features included calcification loads of noncoronary, right coronary, and left coronary cusps (NCC, RCC, and LCC, respectively), aortic annulus area, sinus of Valsalva (SOV) diameter at NCC, sinotubular junction (STJ) diameter (long-axis), ascending aorta diameter (long-axis), right coronary artery (RCA) ostial height, left main coronary artery (LMCA) ostial height, SOV height at NCC, SOV height at RCC, and aortic root angle (**Figure S2B and S2C**). All features were measured in diastole except for the calcification loads of different valve leaflets and aortic annulus area, which were measured in systole. Additionally, three features previously associated with CDs—infra-annular membranous septal length^10^ and left ventricular outflow tract (LVOT) calcification^11^ were assessed as previously described and included in multivariable logistic regression analysis of CDs as covariates.

### AHC of CTA features

AHC of the male and female datasets was separately performed using RStudio (R version 4.0.2) because nearly all CTA features examined herein showed significant sex-specific differences (**Figure S3**). For each dataset, the data variables (i.e., nonredundant CTA features) were prepared by transformation to normality as needed. Following standardization of all variables, the data were assessed for cluster tendency using Hopkins statistic (via the get_cluster_tendency function). AHC was then performed on the dataset with different types of linkage methods (complete, single, average, Ward’s) assuming Euclidean distance, with agglomerative coefficients assessed (via the hcluster and agnes functions). The optimal number of clusters for each dataset was determined based on 30 previously validated indices (via the NbClust function/package).^12^

### Implantation depth measurement

The aortogram obtained during TAVR for each patient was retrospectively reviewed using either IntelliSpace PACS Enterprise (version 4.4.544.0, Philips; VAPAHCS) or IntelliSpace Cardiovascular (version 3.2, Philips, Stanford University). Implantation depth was measured in the right anterior oblique projection as the arithmetic mean of two measured distances: 1) distance between NCC and the distal part of THV; 2) distance between LCC and the distal part of THV.^13^ Each measurement on-screen was calibrated against the known external diameter of the pigtail catheter used to obtain the aortogram.

### Statistical analyses

All statistical analyses were performed using a combination of GraphPad Prism (v6.01, GraphPad Software) and StataNow (v18 BE, StataCorp LLC). Data were presented as mean±SD for normally distributed variables, median (interquartile range) for non-normally distributed variables, and frequency for categorical variables. The normality of a continuous variable was tested using the skewness and kurtosis test. Student’s t-test was used to compare the means of a normally distributed variable between two independent groups, whereas Wilcoxon rank-sum test was used to compare the medians of a non-normally distributed variable between two independent groups. Fisher’s exact test was used to compare the frequency of a categorical variable between two independent groups. One-way repeated measures ANOVA followed by post hoc pairwise paired t-tests were used to compare the means of normally distributed variables across multiple related groups. Friedman test followed by post hoc pairwise Wilcoxon signed-rank tests were used to compare the mean ranks of non-normally distributed variables across multiple related groups. Multiple logistic regression analysis was conducted to determine the association between TAVR-related CDs (a composite endpoint including new LBBB, transient CDs [LBBB or HAVB], or HAVB leading to permanent pacemaker implantation during or within 30 days of TAVR) and AHC-determined cluster type (predictor variable), with covariates selected according to methods further described in **Supplemental Methods.** The likelihood-ratio test was used to assess the incremental prognostic value of cluster type by comparing the goodness-of-fit between the full multivariable logistic regression model and a nested model of the same covariates but without cluster type as an independent variable. Sample size for the study was determined following recommended guidelines^14,15^ as further described in **Supplemental Methods**. *P* value < 0.05 was generally considered statistically significant. *P_adj_* represents the *P* value adjusted for multiple comparisons using Bonferroni correction.

## RESULTS

### Clinical profiles of the study cohort

The clinical characteristics of the study patients are shown in **Table 1**. By study design, the patients are equally divided by sex, with male patients slightly younger (79.0 [72.5, 85.0] versus 83.0 [77.0, 88.0] years old, *P*<0.001) and having greater body mass index (BMI; 27.6 [25.0, 31.7] versus 26.5 [22.5, 31.3] kg/m^2^, *P*=0.001). Compared to female patients, male patients had greater prevalence of some cardiovascular risk factors, including dyslipidemia (80.6% versus 63.0%, *P*<0.001), chronic kidney disease (33.3% versus 24.0%, *P*=0.010), active smoking (18.2% versus 3.3%, *P*<0.001) and comorbidities, including coronary artery disease (75.8% versus 45.0%, *P*<0.001) and left ventricular systolic dysfunction (16.1% versus 6.7%, *P*<0.001). However, they presented with less aortic valve mean gradient (44.0 [40.0, 51.0] versus 46.0 [40.0, 58.0] mmHg, *P*=0.004) and greater aortic valve area (0.77 [0.64, 0.88] versus 0.66 [0.56, 0.74] cm^2^, *P*<0.001) at the time of TAVR evaluation. Regarding risk factors of CDs, while male patients had greater prevalence of pre-existing RBBB (14.2% versus 7.3%, *P*=0.005), females had greater prevalence of LVOT calcification (47.0% versus 38.2%, *P*=0.027). No significant differences in membranous septal length (2.75 [1.59, 4.26] versus 3.10 [1.50, 4.84] mm, *P*=0.122) or implantation depth (5.91 [4.18, 8.73] versus 6.17 [4.38, 9.55] mm, *P*=0.297) were observed between the two groups. Additionally, male patients were associated with greater prevalence of the composite endpoint of TAVR-related CDs (18.5% versus 6.4%, *P*<0.001).

**Table 1.** Clinical profiles of the study cohort stratified by sex. Abbreviations: AS, aortic stenosis; AV, atrioventricular; CVA, cerebrovascular accident; LV, left ventricular; LVOT, left ventricular outflow tract; MG, mean gradient; TIA, transient ischemic attack. *, statistical significance (*P*<0.05).

| Clinical Variable | Male<br>(N=330) | Female<br>(N=330) | *P* Value |
| --- | --- | --- | --- |
| **Basic demographics** |  |  |  |
| Age, years | 79.0 (72.5, 85.0) | 83.0 (77.0, 88.0) | <0.001* |
| Body mass index, kg/m<sup>2</sup> | 27.6 (25.0, 31.7) | 26.5 (22.5, 31.3) | 0.001* |
| **Clinical risk factors, n (%)** |  |  |  |
| Hypertension | 285 (86.4) | 292 (88.5) | 0.481 |
| Dyslipidemia | 266 (80.6) | 208 (63.0) | <0.001* |
| Diabetes | 126 (38.2) | 102 (30.9) | 0.060 |
| Chronic kidney disease | 110 (33.3) | 79 (24.0) | 0.010* |
| Active smoking | 60 (18.2) | 11 (3.3) | <0.001* |
| **Comorbidities, n (%)** |  |  |  |
| Coronary artery disease | 250 (75.8) | 148 (45.0) | <0.001* |
| LV systolic dysfunction | 53 (16.1) | 22 (6.7) | <0.001* |
| Atrial fibrillation/flutter | 75 (22.7) | 64 (19.4) | 0.340 |
| Previous TIA/CVA | 66 (20.0) | 76 (23.0) | 0.394 |
| Peripheral artery disease | 42 (12.7) | 55 (16.7) | 0.187 |
| **AS severity** |  |  |  |
| High-gradient AS, n (%) | 251 (76.1) | 275 (83.3) | 0.026* |
| Low-gradient AS, n (%) | 74 (22.4) | 53 (16.1) | 0.048* |
| Moderate AS, n (%) | 4 (1.2) | 2 (0.6) | 0.686 |
| Aortic valve MG, mmHg | 44.0 (40.0, 51.0) | 46.0 (40.0, 58.0) | 0.004* |
| Aortic valve area, cm<sup>2</sup> | 0.77 (0.64, 0.88) | 0.66 (0.56, 0.74) | < 0.001* |
| **Risk factors for conduction disturbances** |  |  |  |
| First-degree AV block, n (%) | 97 (29.4) | 67 (20.3) | 0.009* |
| Right bundle branch block, n (%) | 47 (14.2) | 24 (7.3) | 0.005* |
| LVOT calcification, n (%) | 126 (38.2) | 155 (47.0) | 0.027* |
| Membranous septal length, mm | 2.75 (1.59, 4.26) | 3.10 (1.50, 4.84) | 0.122 |
| Implantation depth, mm | 5.91 (4.18, 8.73) | 6.17 (4.38, 9.55) | 0.297 |

| **TAVR-related conduction disturbances, n (%)** |  |  |  |
| --- | --- | --- | --- |
| New left bundle branch block | 4 (1.2) | 9 (2.7) | 0.262 |
| Transient conduction disturbances | 5 (1.5) | 2 (0.6) | 0.451 |
| High-degree AV block leading to permanent pacemaker implantation | 52 (15.8) | 10 (3.0) | <0.001* |
| Composite of above variables | 61 (18.5) | 21 (6.4) | <0.001* |

### AHC of CTA features

Before performing cluster analysis, we first reduced both male and female pre-TAVR CTA datasets from 21 to 12 features by eliminating redundant features with high degrees of correlation (Pearson correlation coefficient ≥ 0.8; **Figure S2**). We then confirmed the clustering tendency of the male and female datasets by Hopkins statistic (0.661 and 0.644, respectively). When performing AHC of the two datasets using different linkage methods (complex, single, average, and Ward’s), we found the Ward’s linkage method to consistently yield the highest agglomerative coefficient (male: 0.929, **Figure S4A-S4D**; female: 0.934, **Figure S5A-S5D**). Comparing the heatmaps of the AHC performed with Ward’s linkage on the two datasets (**Figure 1A and 1B**), we noticed similar general patterns of clustering for the CTA features. Specifically, the aortic valve leaflet calcification loads tended to cluster more closely with cross-sectional dimensions (e.g., aortic annulus area, SOV/STJ/ascending aorta diameters) than with height dimensions (SOV heights, LMCA/RCA ostial heights). While the heatmap for the male dataset revealed three visually distinct regions of normalized feature intensity, the corresponding heatmap for the female dataset revealed only two such visually distinct regions. Recognizing that there is not a perfect index for determining the optimal number of clusters for any given dataset, we applied 30 previously validated cluster indices (**Table S2**) to assess our datasets and found that the majority of the indices suggested three and two as the optimal number of clusters for the male and female datasets, respectively (**Figure 1C and 1D**).

**Figure 1.**
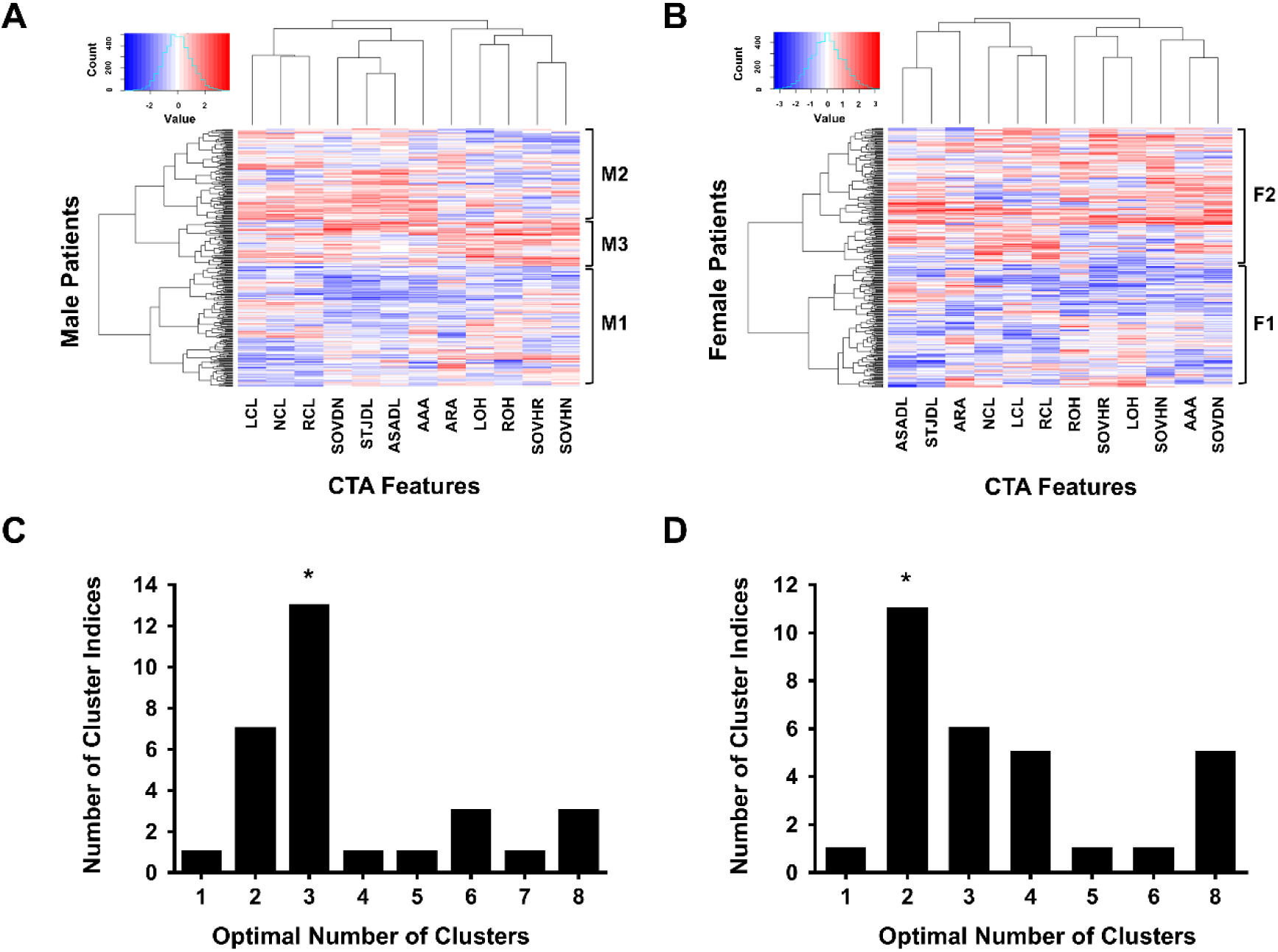
Heatmaps of agglomerative hierarchical clustering and determination of the optimal number of clusters. The top panel shows heatmaps corresponding to agglomerative hierarchical clustering of the (A) male and (B) female datasets, with distinct clusters later identified (M1-M3; F1-F2) indicated. Each column represents a different CTA feature, and each row represents a different patient. The bottom panel shows bar graphs plotting the number of cluster indices (out of 30) that suggested a specific optimal number of clusters for the (C) male and (D) female datasets. *, optimal number of clusters determined, as supported by the most number of cluster indices.

### Clinical characteristics of AHC-derived clusters

Following AHC and cluster membership assignment of the male dataset, we compared the three male clusters identified (M1, M2, and M3) in terms of their clinical characteristics (**Table 2**). The clusters were not significantly different in terms of basic demographics, major cardiovascular risk factors, or common comorbidities, but were different in terms of AS subtypes and hemodynamic parameters. The prevalence of low-gradient AS was significantly higher in M1 than M2 and M3 (29.8% versus 17.1% and 15.3%), with statistical significance between M1 and M2 (*P_adj_*=0.045), but not between M1 and M3 (*P_adj_*=0.075) or between M2 and M3 (*P_adj_*>0.999). The prevalence of high-gradient AS was significantly less in M1 than M2 and M3 (68.8% versus 82.1% and 83.1%), with statistical significance between M1 and M2 (*P_adj_*=0.039) but not between M1 and M3 (*P_adj_*=0.075) or between M2 and M3 (*P_adj_*>0.999). Correspondingly, M1 was associated with a lower aortic valve mean gradient than M2 and M3 (43.0 [33.0, 49.0] versus 45.0 [41.0, 52.0] and 46.0 [40.0, 51.0] mmHg), with significant difference between M1 and M2 (*P_adj_*=0.016) but not between M1 and M3 (*P_adj_*=0.486) or between M2 and M3 (*P_adj_*>0.999). The aortic valve area was not significantly different among the three male clusters (*P_adj_*>0.05 for all pairwise comparisons). None of the previously reported risk factors for TAVR-related CDs (i.e., first-degree atrioventricular [AV] block, RBBB, LVOT calcification, membranous septal length, and implantation depth) were significantly different among the three male clusters (*P*>0.05 for all).

**Table 2.** Patient characteristics of male clusters. Abbreviations: AS, aortic stenosis; AV, atrioventricular; CVA, cerebrovascular accident; LV, left ventricular; LVOT, left ventricular outflow tract; MG, mean gradient; RBBB, right bundle branch block; TIA, transient ischemic attack. *, statistical significance (*P*<0.05).

| Clinical Variable | Cluster M1<br>(N=154) | Cluster M2<br>(N=117) | Cluster M3<br>(N=59) | *P* Value |
| --- | --- | --- | --- | --- |
| **Basic demographics** |  |  |  |  |
| Age, years | 78.5 (71.3, 84.0) | 79.0 (72.0, 86.0) | 82 (74.5, 86.0) | 0.129 |
| Body mass index, kg/m<sup>2</sup> | 27.3 (25.0, 30.7) | 27.9 (25.1, 33.0) | 27.8 (24.5, 31.6) | 0.388 |
| **Clinical risk factors, n (%)** |  |  |  |  |
| Hypertension | 135 (87.7) | 97 (82.9) | 53 (89.8) | 0.366 |
| Dyslipidemia | 128 (83.1) | 90 (76.9) | 48 (81.4) | 0.437 |
| Diabetes | 62 (40.3) | 44 (37.6) | 20 (33.9) | 0.685 |
| Chronic kidney disease | 53 (34.4) | 37 (31.6) | 20 (33.9) | 0.885 |
| Active smoking | 32 (20.8) | 15 (12.8) | 13 (22.0) | 0.170 |
| **Comorbidities, n (%)** |  |  |  |  |
| Coronary artery disease | 120 (77.9) | 84 (71.8) | 46 (78.0) | 0.461 |
| LV systolic dysfunction | 24 (15.6) | 17 (14.5) | 12 (20.3) | 0.597 |
| Atrial fibrillation/flutter | 30 (19.5) | 28 (23.9) | 17 (28.8) | 0.322 |
| Previous TIA/CVA | 27 (17.5) | 25 (21.4) | 14 (23.7) | 0.539 |
| Peripheral artery disease | 19 (12.3) | 11 (9.4) | 12 (20.3) | 0.119 |
| **AS severity** |  |  |  |  |
| High-gradient AS, n (%) | 106 (68.8) | 96 (82.1) | 49 (83.1) | 0.016* |
| Low-gradient AS, n (%) | 46 (29.8) | 20 (17.1) | 9 (15.3) | 0.021* |
| Moderate AS, n (%) | 2 (1.3) | 1 (0.9) | 1 (1.7) | 0.883 |
| Aortic valve MG, mmHg | 43.0 (33.0, 49.0) | 45.0 (41.0, 52.0) | 46.0 (40.0, 50.5) | 0.018* |
| Aortic valve area, cm<sup>2</sup> | 0.77 (0.62, 0.87) | 0.80 (0.70, 0.90) | 0.70 (0.60, 0.80) | 0.050 |
| **Risk factors for conduction disturbances** |  |  |  |  |
| First-degree AV block, n (%) | 38 (24.7) | 36 (30.8) | 23 (39.0) | 0.112 |
| RBBB, n (%) | 18 (11.7) | 23 (19.7) | 6 (10.2) | 0.109 |
| LVOT calcification, n (%) | 51 (33.1) | 49 (41.9) | 26 (44.1) | 0.200 |
| Membranous septal length, mm | 3.06 (1.86, 4.61) | 2.59 (1.63, 4.02) | 2.71 (0.22, 4.15) | 0.135 |
| Implantation depth, mm | 5.755 (4.215, 7.713) | 6.190 (3.865, 9.020) | 6.530 (4.710, 9.980) | 0.366 |

Following AHC of the female dataset and cluster membership assignment, we compared the clinical characteristics of the two female clusters identified (F1 and F2; **Table 3**). We found the two clusters to be similar in basic demographics, major cardiovascular risk factors, and common comorbidities, except for a greater prevalence of peripheral artery disease in F1 compared to F2 (23.1% versus 10.9%, *P*=0.005). Compared to F1, F2 was associated with a higher aortic valve mean gradient (47.5 [41.0, 60.0] versus 44.4 [40.0, 53.0] mmHg, *P*=0.014) and prevalence of LVOT calcification (56.3% versus 36.5%, *P*<0.001).

**Table 3.** Patient characteristics of female clusters. Abbreviations: AS, aortic stenosis; AV, atrioventricular; CVA, cerebrovascular accident; MG, mean gradient; LV, left ventricular; LVOT, left ventricular outflow tract; RBBB, right bundle branch block; TIA, transient ischemic attack. *, statistical significance (*P*<0.05).

| Clinical Variable | Cluster F1<br>(N=156) | Cluster F2<br>(N=174) | *P* Value |
| --- | --- | --- | --- |
| **Basic demographics** |  |  |  |
| Age, years | 83.0 (77.0, 87.3) | 83.0 (78.0, 88.0) | 0.417 |
| Body mass index, kg/m<sup>2</sup> | 26.3 (22.8, 30.6) | 26.8 (22.3, 31.8) | 0.640 |
| **Clinical risk factors, n (%)** |  |  |  |
| Hypertension | 142 (91.0) | 150 (86.2) | 0.226 |
| Dyslipidemia | 102 (65.4) | 106 (60.9) | 0.790 |
| Diabetes | 52 (33.3) | 50 (28.7) | 0.404 |
| Chronic kidney disease | 41 (26.5) | 38 (21.8) | 0.368 |
| Active smoking | 6 (3.8) | 5 (2.9) | 0.762 |
| **Comorbidities, n (%)** |  |  |  |
| Coronary artery disease | 77 (49.7) | 71 (40.8) | 0.123 |
| LV systolic dysfunction | 12 (8) | 10 (5.7) | 0.514 |
| Atrial fibrillation/flutter | 28 (17.9) | 36 (20.7) | 0.578 |
| Previous TIA/CVA | 36 (23.1) | 40 (23.0) | 1.000 |
| Peripheral artery disease | 36 (23.1) | 19 (10.9) | 0.005* |
| **AS severity** |  |  |  |
| High-gradient AS, n (%) | 125 (80.1) | 150 (86.2) | 0.143 |
| Low-gradient AS, n (%) | 29 (18.6) | 24 (13.8) | 0.293 |
| Moderate AS, n (%) | 2 (1.3) | 0 (0) | 0.223 |
| Aortic valve MG, mmHg | 44.4 (40.0, 53.0) | 47.5 (41.0, 60.0) | 0.014* |
| Aortic valve area, cm<sup>2</sup> | 0.68 (0.59, 0.74) | 0.66 (0.55, 0.72) | 0.250 |
| **Risk factors for conduction disturbances** |  |  |  |
| First-degree AV block, n (%) | 38 (24.4) | 29 (16.7) | 0.100 |
| RBBB, n (%) | 8 (5.1) | 16 (9.2) | 0.203 |
| LVOT calcification, n (%) | 57 (36.5) | 98 (56.3) | <0.001* |
| Membranous septal length, mm | 3.07 ± 0.19 | 3.15 ± 0.20 | 0.778 |
| Implantation depth, mm | 6.77 (4.51, 9.55) | 5.77 (4.13, 9.66) | 0.307 |

### CTA features of AHC-derived clusters

The male and female clusters could be readily distinguished by their CTA features (**Tables 4 and 5**). Notably, all 12 CTA features were significantly different among the three male clusters (all *P*<0.001 in **Table 4**; **Table S3** for pairwise comparisons). Compared to both M2 and M3, M1 had smaller calcification loads for all three leaflets: NCC (304.4 [220.3, 422.3] versus 508.0 [333.6, 739.0] and 485.0 [327.0, 694.5] AU, both *P_ad_j*<0.001), RCC (200.7 [118.6, 330.6] versus 407.0 [276.0, 594.1] and 332.0 [230.2, 514.5] AU, both *P_adj_*<0.001), and LCC (186.5 [110.5, 351.9] versus 378.0 [264.0, 547.0] and 293.0 [190.0, 457.5] AU, *P_adj_*<0.001 and *P_adj_*=0.002, respectively). M1 also had relatively smaller aortic root angle (45.0±8.7 versus 47.6±6.7 and 52.0±7.6 degrees, *P_adj_*=0.016 and *P_adj_*<0.001, respectively) and cross-sectional aortic root dimensions than M2 and M3, as exemplified by smaller aortic annulus area (4.89±0.63 versus 5.34±0.63 and 5.46±0.65 cm^2^, both *P_adj_*<0.001), SOV diameter at NCC (32.8±2.3 versus 35.1±2.2 and 36.9±2.7 mm, *P_adj_*<0.001 for both), STJ diameter (29.6 [28.0, 30.9] versus 32.7 [31.2, 34.8] and 31.9 [30.8, 34.0] mm, both *P_adj_*<0.001), and ascending aorta diameter (32.8 [31.4, 34.0] versus 36.1 [34.3, 38.0] and 34.6 [33.7, 36.7] mm, both *P_adj_*<0.001). The most noticeable differences between M2 and M3 were the smaller aortic root height dimensions of M2, as evidenced by smaller RCA ostial height (17.4±2.7 versus 21.3±3.2 mm, *P_adj_*<0.001), LMCA ostial height (14.7±2.4 versus 17.9±2.4 mm, *P_adj_*<0.001), SOV height at NCC (22.5 [20.2, 25.0] versus 28.0 [25.3, 29.8] mm, *P_adj_*<0.001), and SOV height at RCC (20.1±4.6 versus 27.3±5.0 mm, *P_adj_*<0.001).

**Table 4.** Computed tomography angiography features of male clusters. Abbreviations: CTA, computed tomography angiography; LCC, left coronary cusp; LMCA, left main coronary artery; NCC, noncoronary cusp; RCA, right coronary artery; RCC, right coronary cusp; SOV, sinus of Valsalva; STJ, sinotubular junction. *, statistical significance (*P*<0.05) comparing all three clusters.

| CTA Feature | Cluster M1<br>(N=154) | Cluster M2<br>(N=117) | Cluster M3<br>(N=59) | P Value |
| --- | --- | --- | --- | --- |
| NCC calcification load, AU | 304.4 (220.3, 422.3) | 508.0 (333.6, 739.0) | 485.0 (327.0, 694.5) | <0.001* |
| RCC calcification load, AU | 200.7 (118.6, 330.6) | 407.0 (276.0, 594.1) | 332.0 (230.2, 514.5) | <0.001* |
| LCC calcification load, AU | 186.5 (110.5, 351.9) | 378.0 (264.0, 547.0) | 293.0 (190.0, 457.5) | <0.001* |
| Aortic annulus area, cm<sup>2</sup> | 4.89 ± 0.63 | 5.34 ± 0.63 | 5.46 ± 0.65 | <0.001* |
| SOV diameter at NCC, mm | 32.8 ± 2.3 | 35.1 ± 2.2 | 36.9 ± 2.7 | <0.001* |
| STJ diameter, mm | 29.6 (28.0, 30.9) | 32.7 (31.2, 34.8) | 31.9 (30.8, 34.0) | <0.001* |
| Ascending aorta diameter, mm | 32.8 (31.4, 34.0) | 36.1 (34.3, 38.0) | 34.6 (33.7, 36.7) | <0.001* |
| Aortic root angle, degrees | 45.0 ± 8.7 | 47.6 ± 6.7 | 52.0 ± 7.6 | <0.001* |
| RCA ostial height, mm | 17.2 ± 3.0 | 17.4 ± 2.7 | 21.3 ± 3.2 | <0.001* |
| LMCA ostial height, mm | 15.1 ± 2.4 | 14.7 ± 2.4 | 17.9 ± 2.4 | <0.001* |
| SOV height at NCC, mm | 23.4 (21, 25.2) | 22.5 (20.2, 25.0) | 28.0 (25.3, 29.8) | <0.001* |
| SOV height at RCC, mm | 21.8 ± 4.8 | 20.1 ± 4.6 | 27.3 ± 5.0 | <0.001* |

**Table 5.** Computed tomography angiography features of female clusters. Abbreviations: CTA, computed tomography angiography; LCC, left coronary cusp; LMCA, left main coronary artery; NCC, noncoronary cusp; RCA, right coronary artery; RCC, right coronary cusp; SOV, sinus of Valsalva; STJ, sinotubular junction. *, statistical significance (*P*<0.05).

| CTA Feature | Cluster F1<br>(N=156) | Cluster F2<br>(N=174) | *P* Value |
| --- | --- | --- | --- |
| NCC calcification load, AU | 242.0 (153.8, 350.5) | 441.5 (290.0, 647.0) | <0.001* |
| RCC calcification load, AU | 127.0 (77.7, 214.0) | 287.0 (212.3, 418.8) | <0.001* |
| LCC calcification load, AU | 138.0 (82.2, 218.3) | 261.0 (174.3, 418.5) | <0.001* |
| Aortic annulus area, cm<sup>2</sup> | 3.57 ± 0.04 | 4.14 ± 0.04 | <0.001* |
| SOV diameter at NCC, mm | 28.0 ± 1.8 | 30.7 ± 2.3 | <0.001* |
| STJ diameter, mm | 25.6 (23.8, 26.9) | 27.7 (26.5, 29.9) | <0.001* |
| Ascending aorta diameter, mm | 30.8 (28.9, 32.4) | 32.3 (30.8, 34.6) | <0.001* |
| Aortic root angle, degrees | 45.9 ± 8.6 | 47.3 ± 7.6 | 0.101 |
| RCA ostial height, mm | 13.7 ± 2.9 | 14.9 ± 2.8 | <0.001* |
| LMCA ostial height, mm | 12.6 ± 2.5 | 13.3 ± 2.4 | 0.014* |
| SOV height at NCC, mm | 19.8 (18.3, 21.0) | 22.5 (20.3, 24.1) | <0.001* |
| SOV height at RCC, mm | 20.2 ± 2.9 | 21.8 ± 3.1 | <0.001* |

In contrast to the more complicated feature patterns of male clusters, the CTA features of female clusters were more predictable. All features except for aortic root angle were significantly different between the two female clusters such that their values were always lower in F1 than F2 (**Table 5**).

### TAVR-related CDs among AHC-determined clusters

To determine how TAVR-related CDs as a composite endpoint is related to the AHC-determined clusters, we performed multivariable logistic regression on a combined male/female dataset with TAVR-related CDs as the outcome variable and cluster type as the predictor variable, while adjusting for covariates selected as described in **Supplemental Methods** (**Table 6**). We found cluster type to be a significant predictor of TAVR-related CDs. Compared to M1, M2 was more associated with CDs (odds ratio [OR]=2.150, *P*=0.032), whereas M3 was not significantly different from either M1 (OR=2.120, *P*=0.085) or M2 (OR=0.986, *P*=0.974). The female clusters (F1 and F2) were not significantly different from M1 (OR=0.696, *P*=0.414; OR=0.901, *P*=0.812) or from each other (OR=1.294, *P*=0.581). Consistent with the literature, many of the covariates previously reported to be risk factors for CDs were also found to be significantly associated with CDs, including first-degree AV block (OR=1.872, *P*=0.027), RBBB (OR=2.467, *P*=0.007), membranous septal length (OR=0.880, *P*=0.012), implantation depth (OR=1.073, *P*=0.011), and TAVR THV type (OR=2.923, *P*=0.036 comparing Lotus valve to CoreValve; OR=0.357, *P*<0.001 comparing Edwards SAPIEN 3 valve to CoreValve; OR=0.122, *P*<0.001 comparing Edward SAPIEN 3 valve to Lotus valve). By likelihood-ratio test, the multivariable regression model, including cluster type as a predictor, had significantly better goodness-of-fit than a nested model without cluster type as predictor (*P*=0.020), suggesting the incremental prognostic value of AHC-determined cluster type.

**Table 6.**
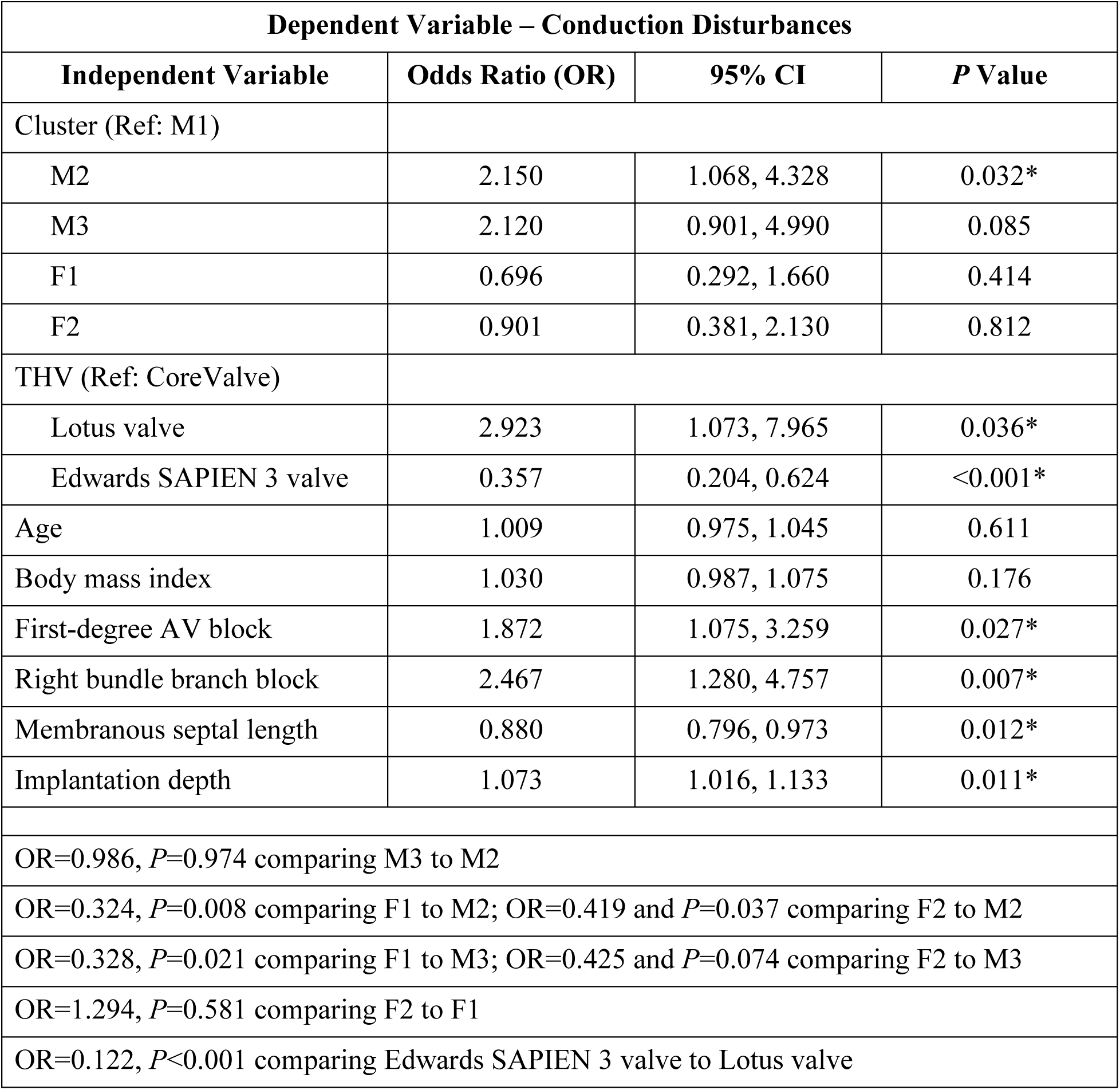
Summary of logistic regression analysis on conduction disturbances. Abbreviations: AV, atrioventricular; F, female cluster; M, male cluster. *, statistical significance (*P*<0.05).

| Dependent Variable – Conduction Disturbances |  |  |  |
| --- | --- | --- | --- |
| Independent Variable | Odds Ratio (OR) | 95% CI | P Value |
| Cluster (Ref: M1) |  |  |  |
| M2 | 2.150 | 1.068, 4.328 | 0.032* |
| M3 | 2.120 | 0.901, 4.990 | 0.085 |
| F1 | 0.696 | 0.292, 1.660 | 0.414 |
| F2 | 0.901 | 0.381, 2.130 | 0.812 |
| THV (Ref: CoreValve) |  |  |  |
| Lotus valve | 2.923 | 1.073, 7.965 | 0.036* |
| Edwards SAPIEN 3 valve | 0.357 | 0.204, 0.624 | <0.001* |
| Age | 1.009 | 0.975, 1.045 | 0.611 |
| Body mass index | 1.030 | 0.987, 1.075 | 0.176 |
| First-degree AV block | 1.872 | 1.075, 3.259 | 0.027* |
| Right bundle branch block | 2.467 | 1.280, 4.757 | 0.007* |
| Membranous septal length | 0.880 | 0.796, 0.973 | 0.012* |
| Implantation depth | 1.073 | 1.016, 1.133 | 0.011* |
| OR=0.986, *P*=0.974 comparing M3 to M2 |  |  |  |
| OR=0.324, *P*=0.008 comparing F1 to M2; OR=0.419 and *P*=0.037 comparing F2 to M2 |  |  |  |
| OR=0.328, *P*=0.021 comparing F1 to M3; OR=0.425 and *P*=0.074 comparing F2 to M3 |  |  |  |
| OR=1.294, *P*=0.581 comparing F2 to F1 |  |  |  |
| OR=0.122, *P*<0.001 comparing Edwards SAPIEN 3 valve to Lotus valve |  |  |  |

## DISCUSSION

The main findings of this study were: 1) In both male and female patients with clinically significant AS undergoing TAVR, the 12 common features of their preoperative CTAs were clusterable based on Hopkins statistic; 2) UML/AHC of these CTA features revealed three and two distinct clusters for the male and female patients, respectively, as supported by evaluation with 30 cluster indices; 3) The clusters associated with each sex type for the most part were not readily distinguishable amongst each other based on demographics, risk factors, or comorbidities; 4) However, they were easily distinguishable based on CTA features; 5) TAVR-related CDs were independently associated with cluster type, after adjusting for age, BMI, THV type, and other known risk factors for CDs; 6) The male cluster associated with the least TAVR-related CDs (M1; reference) was associated with both small aortic valve leaflet calcification loads and aortic root dimensions (i.e., both cross-sectional and height dimensions) and highest prevalence of low-gradient AS, whereas the male cluster associated with most TAVR-related CDs (M2) was associated with large aortic valve leaflet calcification loads, large cross-sectional dimensions of the aortic root, small height dimension of the aortic root (i.e., “short and wide aortic root”), and higher prevalence of high-gradient AS than M1; 7) The female clusters were not significantly different from M1 or from each other in terms of association with CDs; 8) AHC-determined clusters provide incremental prognostic value over many conventional risk factors of CDs. Overall, these results suggest that aortic valve leaflet calcification loads and aortic root/ascending aorta anatomy are collectively important determinants and prognosticators of TAVR-related CDs.

CDs continue to be a serious perioperative complication despite advances in TAVR technology, THV valve design, and methods to preoperatively identify risk factors. Numerous risk factors of CDs have been previously identified, with several of them also found to be significant predictors of CDs in our study: first-degree AV block, RBBB, membranous septal length, and implantation depth, and THV type. While many of the previous studies focused on investigating single or a limited number of risk factors for CDs, this is the first study using UML/AHC to define clusters based on a myriad of CTA imaging features, followed by examining cluster-specific susceptibilities to CDs. The successfully identified clusters represent unique patient phenotypes based on aortic valve leaflet calcification loads and aortic root dimensions on pre-TAVR CTA. Leveraging the strength of cluster analysis to uncover hidden interrelationships among myriads of features, we found that patients with greater aortic valve leaflet calcification generally have wider aortic roots. However, in male but not female patients, those with large aortic valve leaflet calcification loads and wide aortic roots segregate into two subgroups, one with short (M2) and another with tall (M3) aortic roots. These different “shape” subphenotypes break away from the designation of simply “small” or “large” aortic root and allow a more refined examination of their association with CDs.

Because the patient clusters and their separations were derived from AHC of 12 common CTA features, it is assumed that these features collectively contribute to cluster-dependent differences in TAVR-related CDs. Indeed, many of the features examined had been previously implicated in CDs, including most notably, the calcification load of LCC, which is thought to cause shifting of THV towards NCC and RCC during TAVR, thus increasing the likelihood of damage to the nearby conduction system.^6^ However, the calcification loads of other leaflets (also confirmed herein), LVOT and aortomitral continuity have all been reported as predictors of CDs,^16^ suggesting the general detrimental effects of calcification and its asymmetric deposition near THV implantation. The relationships between aortic root dimensions and CDs are relatively more complicated. On the one hand, large prosthetic valves, selected for cases of large aortic annulus, have been shown to increase CD risks, with mechanisms partly thought to be related to device-specific factors.^17^ On the other hand, oversizing of prosthetic valves tends to be greater when the aortic annulus size is smaller,^18^ thus posing increased risks for CDs. Other CTA features used to construct the clusters have also been implicated in CDs to varying degrees, including larger aortic root angle (horizontal aorta),^19^ greater ascending aorta diameter,^20^ and shorter left coronary ostial height.^21^ The last factor implies the need for deeper THV implantation to avoid coronary obstruction and the likelihood of greater LVOT coverage, leading to increased risk of CDs. These implications may also potentially explain why M2 (the male cluster associated with a shorter aortic root than the M3 male cluster), but not M3, was more associated with CDs than M1 (reference). However, because our multivariable logistic regression analysis also adjusted for THV implantation depth, the mechanisms by which a shorter aortic root may predispose to CDs should involve factors beyond its effect on implantation depth. Patient-specific computational simulation of TAVR deployment and its precise mechanical effects on the conduction system should help to further elucidate these factors.^22^

Several studies have previously examined the use of UML for risk stratifying patients undergoing TAVR. For instance, Lachmann et al. used AHC to cluster patients with severe AS based on preprocedural echocardiographic and right heart catheterization data to differentiate those more likely to have poor than favorable survival after TAVR.^23^ Kusunose et al. used k-means clustering to analyze clinical, lab, and echocardiographic data of patients with severe AS to identify patient clusters associated with different outcomes (major adverse cardiovascular events and all-cause mortality) after TAVR.^24^ Meredith et al. also used k-means clustering to cluster patients with severe AS based on demographic, biochemical, and cardiac imaging data and showed that the clusters identified were differentially associated with 12-month mortality after TAVR.^25^ Adding to these foundational works, our study here further demonstrates the feasibility of using AHC of pre-TAVR CTA features to identify patient subphenotypes with differential occurrences of CDs. Importantly, the cluster assignment was found to provide incremental prognostic value over many existing clinical risk factors. Thus, the results here support further efforts to incorporate UML algorithms into pre-TAVR CTAs for enhanced risk stratification of CDs.

## STUDY LIMITATIONS

This study has certain limitations that may affect data interpretation. First, as evidenced in our study, the optimal number of clusters in a dataset may vary depending on the index used to evaluate it. Thus, we applied 30 previously validated cluster indices to derive the optimal numbers of clusters for our datasets following the “majority rule.” As patients with AS are known to be highly heterogenous, the clustering structure may be affected by regional patient characteristics and will warrant further validation at other institutions beyond the two nearby institutions involved in this study. Second, in contrast to the male clusters, the female clusters identified herein were not associated with differential occurrences of CDs after TAVR. However, because the overall prevalence of CDs for female patients was low in this study (6.4%; 3% requiring permanent pacemaker), a larger sample size likely will be needed to discern if there are truly differences in CDs between the two female clusters. Third, this study was powered to assess a composite endpoint of CDs designed to increase the sensitivity of detecting differences among the patient clusters given the study’s sample size. A larger-scale study will thus be needed to instill sufficient power to further delineate how cluster assignment affects individual components of the composite endpoint, which may have different impact on patients’ long-term outcomes.^26^

## CONCLUSIONS

Preoperative CTAs are an integral part of pre-TAVR assessment, but their utilization in risk stratification of TAVR-related CDs have been underappreciated or largely limited to a handful of CTA features, the aggregate effects of which are often difficult to assess. UML/AHC enables the discovery of previously unknown patient subphenotypes (clusters) based on 12 common CTA features, which importantly yield incremental prognostic information on CDs over various existing risk factors in combination. These results herein advocate for a new approach to interpreting pre-TAVR CTAs for the assessment of CD risks, in which the focus lies in identifying specific subphenotypes of unique risk profiles, rather than considering the additive risks from selected features. With further validation and integration into the current clinical workflow of pre-TAVR CTAs (e.g., with automated feature extraction), UML can help physicians more efficiently and accurately predict TAVR-related CD risks so that proper risk mitigation strategies (e.g., THV section, depth/approach of implantation) can be planned accordingly to make TAVR safer.

## ACKNOWLEDGEMENTS

This material is the result of work supported with the resources and use of facilities at VAPAHCS. The contents do not represent the views of the Department of Veterans Affairs or the United States Government. We thank Dr. Pei-Yu Lee for her assistance with manuscript preparation.

## SOURCES OF FUNDING

This work was supported in part by the Veterans Affairs Palo Alto Health Care System Administration Office Funding Opportunity Announcement (IYC) and the Stanford Translational Research and Applied Medicine Pilot Grant (IYC).

## DISCLOSURES

None

## SUPPLEMENTAL MATERIALS

Supplemental Methods

Tables S1-S3

Figures S1-S5

## Notes

### Competing Interest Statement

The authors have declared no competing interest.

### Author Declarations

IRB of Stanford University gave ethical approval for this work

