## Supplemental Materials for "Unsupervised Machine Learning of Computed Tomography Angiography Features Uncovers Unique Subphenotypes of Aortic Stenosis With Differential Risks of Conduction Disturbances Following Transcatheter Aortic Valve Replacement"

### SUPPLEMENTAL METHODS

#### Variable selection for logistic regression

In this study, we constructed a multivariable logistic regression model to specifically assess the association between transcatheter aortic valve replacement (TAVR)-related conduction disturbances (CDs; outcome variable) and agglomerative hierarchical clustering (AHC)-determined cluster type (predictor variable). We first forced age and body mass index as essential covariates into the regression model. We then included an additional set of covariates chosen *a priori* based on the literature or their clinical likelihood to influence the relationship between the outcome variable and the predictor variable: transcatheter heart valve type, first-degree atrioventricular block, left bundle branch block, right bundle branch block, atrial fibrillation, implantation depth, left ventricular outflow tract calcification, left ventricular systolic dysfunction, and institution. Some of these variables were further eliminated using an automated stepwise backward selection algorithm, which eliminated covariates that failed to reach a predefined level of statistical significance ($P<0.1$). Sex was not included as a covariate due to its collinearity with cluster type, which was already sex-specific.

#### Sample size determination for the study cohorts

The minimum number of samples needed for this study was based on two main considerations: 1) adequate number of samples needed to form stable clusters by hierarchical clustering of separate male and female datasets; 2) adequate number of samples (in the combined male/female dataset) to allow precise logistic regression analysis to investigate the association between CDs (outcome variable) and cluster type (predictor variable). For efficient clustering, we determined a minimal sample size of 180 (n=90 per male/female dataset) based on a previous recommendation of at least

30 samples per subgroup[14] and an estimated number of subgroups per male/female dataset to be at most three. For sufficient multiple logistic regression analysis, we determined the minimal sample size to be 593, following the guideline of at least 500 samples for multiple regression analysis in observation studies and the events per variable formula of $N = 10 * k/p$ , where $N$ is the minimally required sample size, $k$ the number of independent variables (n=8), and $p$ the proportion of outcome events (estimated to be 13.5% from a prior pilot study of 100 patients).[15] After rounding off to the nearest 10 and adding an additional 10% safety factor to ensure stability of analysis, we eventually implemented a sample size of 660 (n=330 each for the male and female datasets).

| Inclusion Criteria | Exclusion Criteria |
|---|---|
| Diagnosis of calcific (degenerative) aortic stenosis | Bicuspid aortic valve |
| Underwent pre-TAVR CTA | History of prosthetic valve |
| | History of prior mediastinal radiation |
| | History of endocarditis |
| | History of rheumatic valve disease |
| | Inadequate CTA image quality |

**Table S1. Inclusion and exclusion criteria for patient enrollment.** Abbreviations: CTA, computed tomography angiography; TAVR, transcatheter aortic valve replacement.

| | Male Dataset | Female Dataset |
|---|---|---|
| **Index** | **Optimal Number of Clusters Indicated** | **Optimal Number of Clusters Indicated** |
| 1. KL (Krzanowski, Lai) | 6 | 2 |
| 2. CH (Calinski, Harabasz) | 2 | 2 |
| 3. Hartigan (Hartigan) | 3 | 4 |
| 4. CCC (Sarle) | 2 | 2 |
| 5. Scott (Scott, Symons) | 3 | 3 |
| 6. Marriot (Marriot) | 6 | 4 |
| 7. TrCovW (Milligan, Cooper) | 3 | 3 |
| 8. TraceW (Milligan, Cooper) | 3 | 4 |
| 9. Friedman (Friedman, Rubin) | 3 | 3 |
| 10. Rubin (Friedman, Rubin) | 3 | 4 |
| 11. Cindex (Hubert, Levin) | 2 | 2 |
| 12. DB (Davies, Bouldin) | 5 | 8 |
| 13. Silhouette (Rousseeuw) | 2 | 2 |
| 14. Duda (Duda, Hart) | 3 | 2 |
| 15. Pseudot2 (Duda, Hart) | 3 | 2 |
| 16. Beale (Beale) | 2 | 2 |
| 17. Ratkowsky (Ratkowsky, Lance) | 3 | 2 |
| 18. Ball (Ball, Hall) | 3 | 3 |
| 19. Ptbiserial (Milligan) | 8 | 8 |
| 20. Gap (Tibshirani et al.) | 2 | 2 |
| 21. Frey (Frey, Van Groenewoud) | 1 | 1 |
| 22. McClain (McClain, Rao) | 2 | 2 |
| 23. Gamma (Baker, Hubert) | 8 | 8 |
| 24. Gplus (Rohlf, Milligan) | 8 | 8 |
| 25. Tau (Rohlf, Milligan) | 3 | 3 |
| 26. Dunn (Dunn) | 4 | 6 |
| 27. Hubert (Hubert, Arabie) | 3 | 5 |
| 28. SDindex (Halkidi et al.) | 3 | 3 |
| 29. Dindex (Lebart et al.) | 6 | 4 |
| 30. SDbw (Halkidi, Vazirgiannis) | 7 | 8 |

**Table S2. Optimal number of clusters determined by 30 cluster indices for the male and female datasets.** The authors/creators of each cluster index are indicated in parenthesis.

| CTA Features | M1 vs M2 $P_{adj}$ Value | M1 vs M3 $P_{adj}$ Value | M2 vs M3 $P_{adj}$ Value | All Clusters $P$ Value |
|---|---|---|---|---|
| NCC calcification load | <0.001* | <0.001* | 1.000 | <0.001† |
| RCC calcification load | <0.001* | <0.001* | 0.157 | <0.001† |
| LCC calcification load | <0.001* | 0.002* | 0.013* | <0.001† |
| Aortic annulus area | <0.001* | <0.001* | 0.759 | <0.001† |
| SOV diameter at NCC | <0.001* | <0.001* | <0.001* | <0.001† |
| STJ diameter | <0.001* | <0.001* | 0.381 | <0.001† |
| Ascending aorta diameter | <0.001* | <0.001* | 0.013* | <0.001† |
| Aortic root angle | 0.016* | <0.001* | 0.002* | <0.001† |
| RCA ostial height | 1.000 | <0.001* | <0.001* | <0.001† |
| LMCA ostial height | 0.388 | <0.001* | <0.001* | <0.001† |
| SOV height at NCC | 0.222 | <0.001* | <0.001* | <0.001† |
| SOV height at RCC | 0.020* | <0.001* | <0.001* | <0.001† |

**Table S3. *P* values associated with multiple group and individual pairwise comparisons of computed tomography angiography features among/between different male clusters.** Abbreviations: CTA, computed tomography angiography; LCC, left coronary cusp; LMCA, left main coronary artery; NCC, noncoronary cusp; RCA, right coronary artery; RCC, right coronary cusp; SOV, sinus of Valsalva; STJ, sinotubular junction. †, statistical significance ($P$<0.05) comparing all clusters. *, statistical significance from pairwise comparison ($P_{adj}$<0.05, where $P_{adj}$ is adjusted for multiple comparisons by Bonferroni correction).

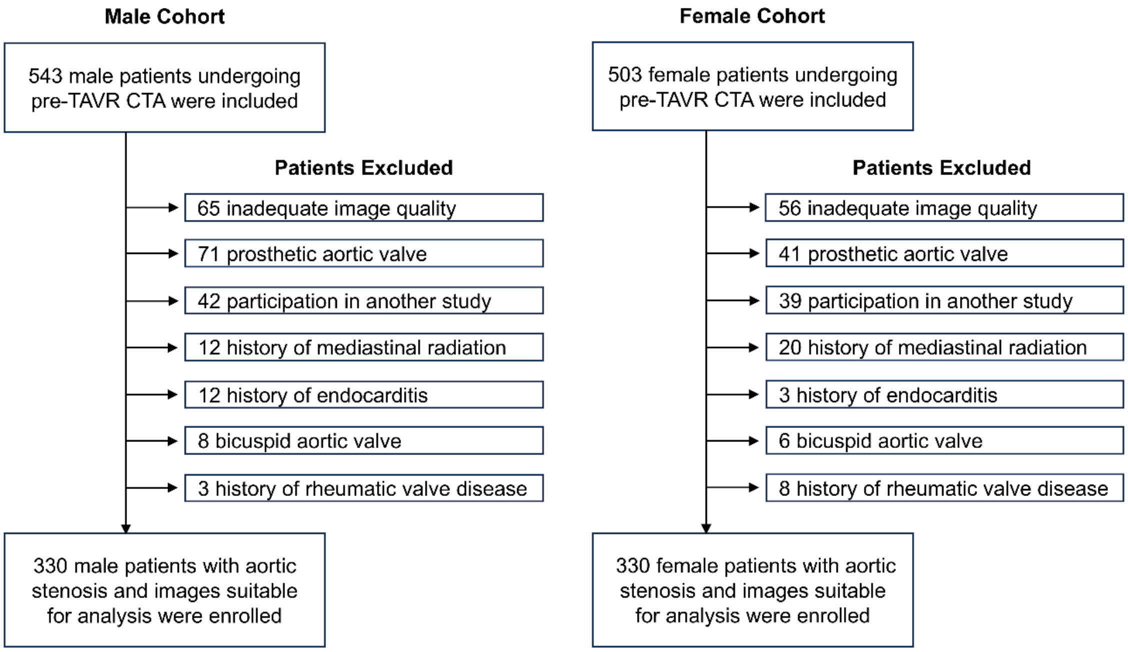


**Figure S1. Flow chart for study enrollment.** Abbreviations: CTA, computed tomography angiography; TAVR, transcatheter aortic valve replacement.

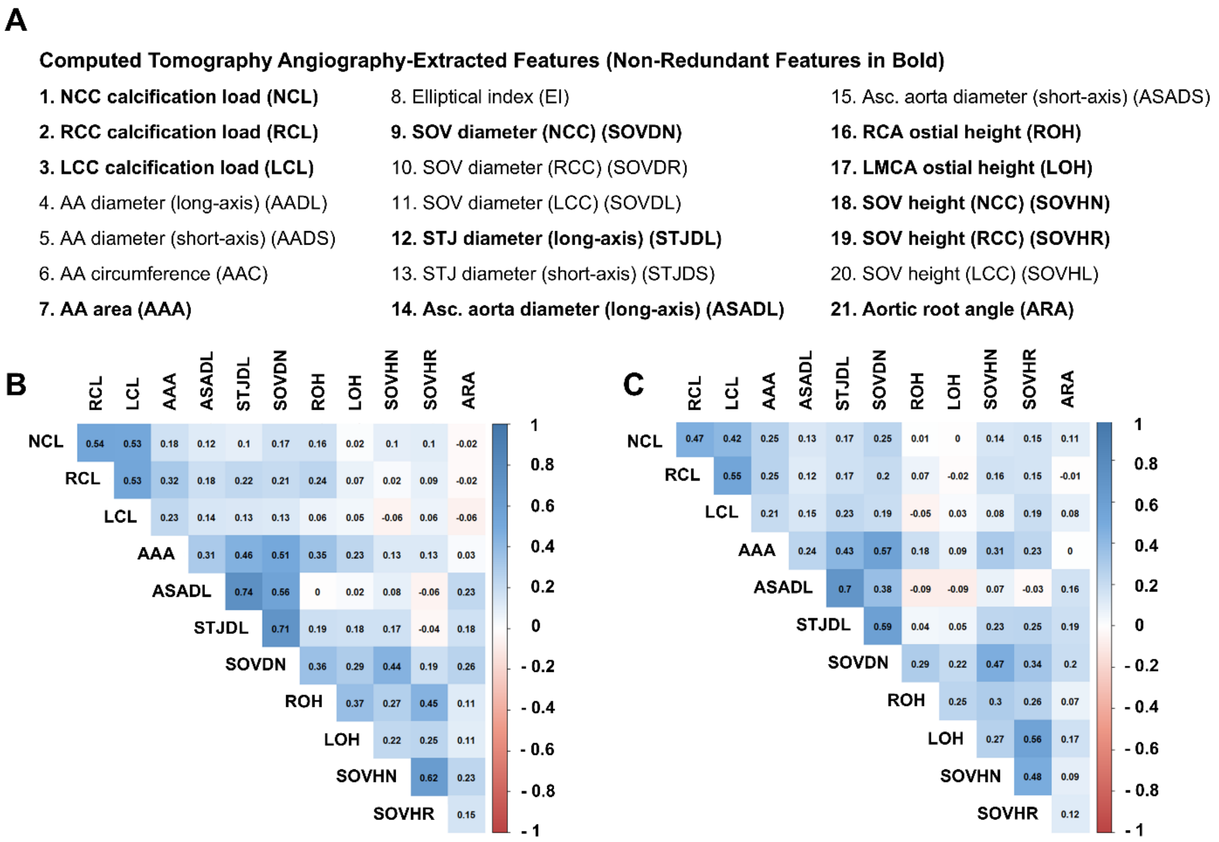


**Figure S2. Computed tomography angiography features extracted and trimmed.** (A) List of imaging features (and their abbreviations in parenthesis) originally extracted from preoperative computed tomography angiography images and later trimmed to a non-redundant set highlighted in bold. (B) The correlation matrix of the final 12 nonredundant features for the male dataset, with Pearson correlation coefficients color-coded and indicated for different pairs of features. (C) The correlation matrix of the final 12 nonredundant features for the female dataset, with Pearson correlation coefficients shown for different pairs of features. Abbreviations: AA, aortic annulus; Asc, ascending; NCC, noncoronary cusp; LCC, left coronary cusp; LMCA, left main coronary artery; RCA, right coronary artery; RCC, right coronary cusp; SOV, sinus of Valsalva; STJ, sinotubular junction.

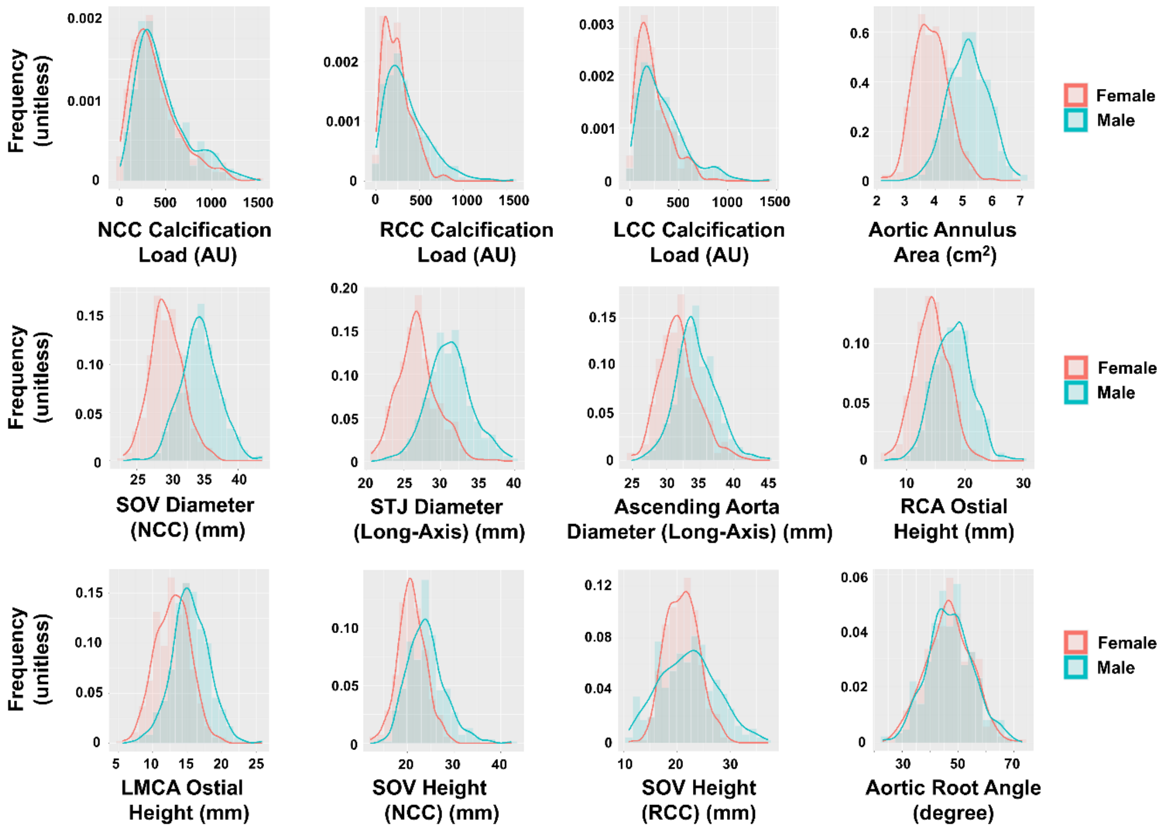


**Figure S3. Frequency histograms of nonredundant computed tomography angiography features.** Frequency histograms of all 12 computed tomography angiography features for the male (turquoise color) and female (salmon color) datasets. Abbreviations: AU, Agatston unit; LCC, left coronary cusp; LMCA, left main coronary artery; NCC, noncoronary cusp; RCA, right coronary artery; RCC, right coronary cusp; SOV, sinus of Valsalva; STJ, sinotubular junction.

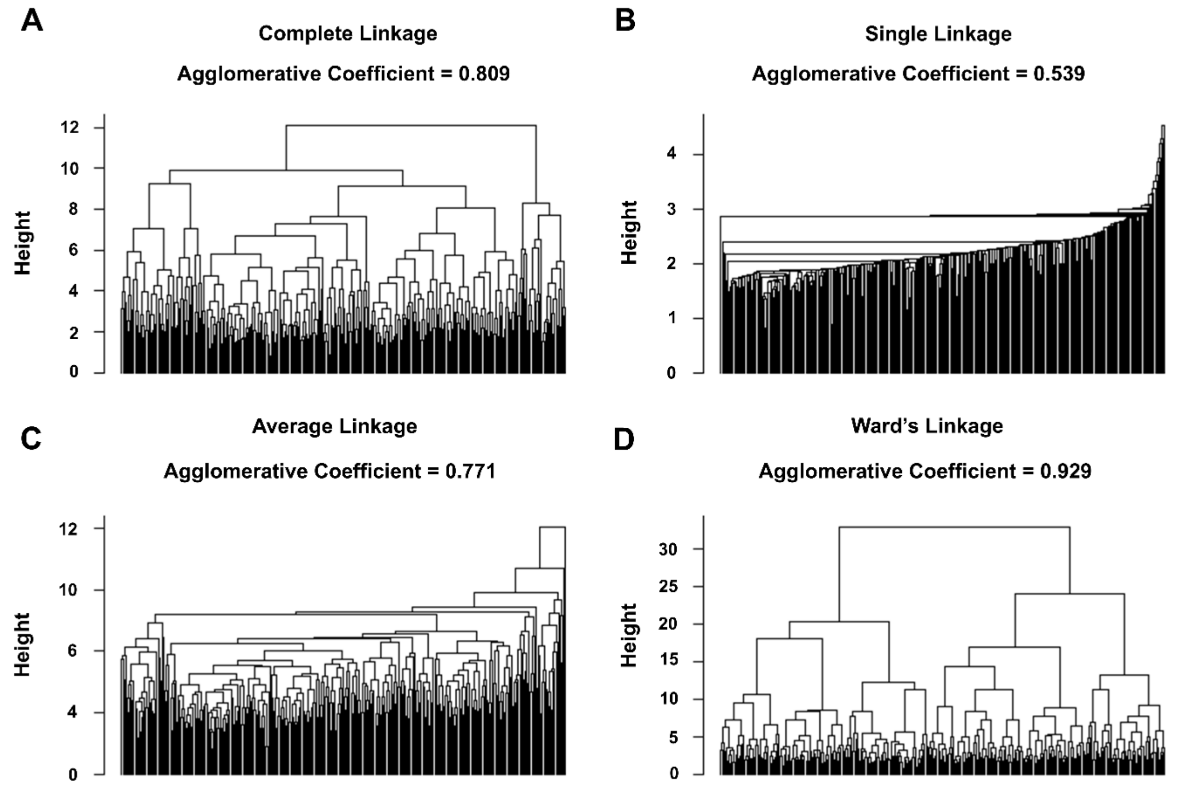


**Figure S4. Dendrograms from agglomerative hierarchical clustering of the male dataset using different linkage methods.** Dendrograms created from agglomerative hierarchical clustering of male datasets using (A) complete linkage, (B) single linkage, (C) average linkage, and (D) Ward's linkage. The agglomerative coefficient achieved with each clustering approach/linkage method is indicated.

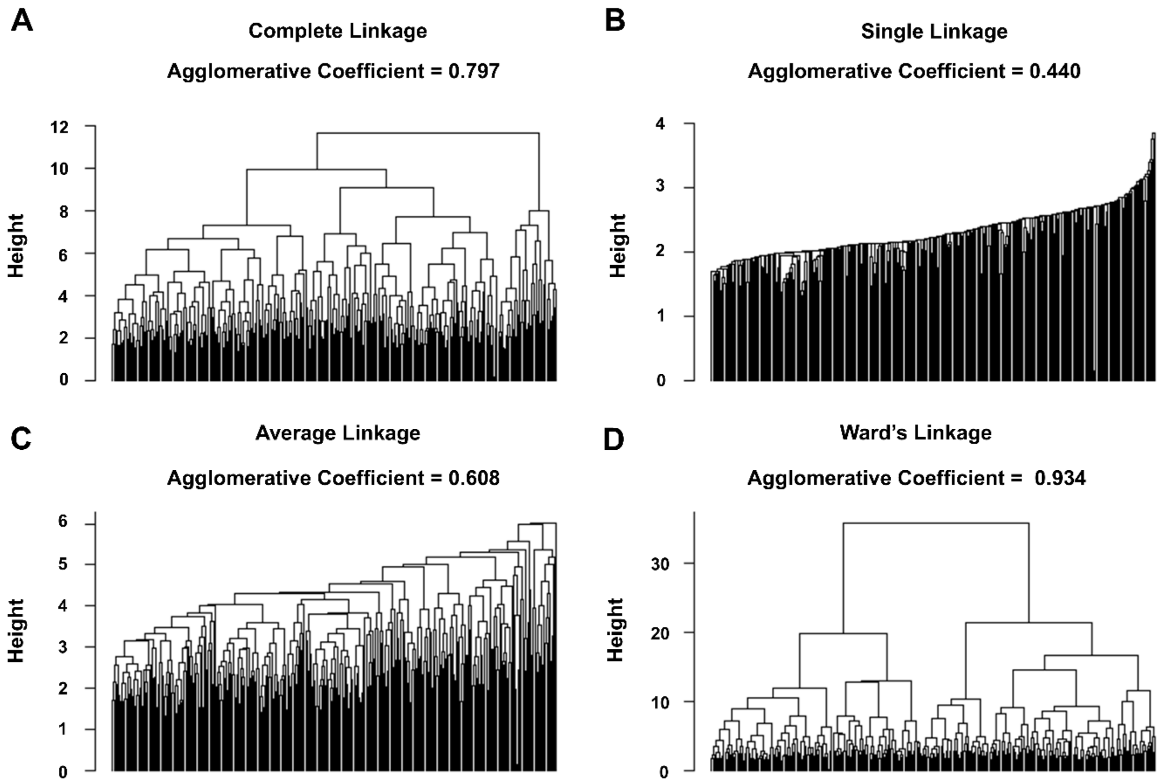


**Figure S5. Dendrograms from agglomerative hierarchical clustering of the female dataset using different linkage methods.** Dendrograms created from agglomerative hierarchical clustering of female datasets using (A) complete linkage, (B) single linkage, (C) average linkage, and (D) Ward's linkage. The agglomerative coefficient achieved with each clustering approach/linkage method is indicated.
